# Clinical study type classification, validation, and PubMed filter comparison with natural language processing and active learning

**DOI:** 10.1101/2022.11.01.22281685

**Authors:** David G P van IJzendoorn, Philippe C Habets, Christiaan H Vinkers, Willem M Otte

**Author notes:** Shared first author (David G P van IJzendoorn, Philippe C Habets). shared last author (Christiaan H Vinkers, Willem M Otte).

## Abstract

Each day, many thousands of new studies are published. Identifying specific study types with high sensitivity and specificity may improve searchability and accelerate updating systematic reviews and meta-analyses. Machine learning transformer models could facilitate this identification process if sufficient training data is available.

We used an active learning strategy to construct a large training set (n=50,000) and fine-tuned the PubMedBERT language model to classify PubMed abstracts as randomized controlled trials, human studies, systematic reviews with and without meta-analyses, protocols, and rodent studies. In an external dataset (n=5,000), the average sensitivity and specificity across study types were 0.94 and 0.96, respectively. PubMed’s internal filters had a low sensitivity for both systematic reviews with meta-analysis (0.175, CI: 0.057–0.293) and randomized controlled trials (0.256, CI: 0.119–0.393). We applied this labeling to all 34 million PubMed abstracts currently available and provide the results within an online meta-information platform (EvidenceHunt).

In conclusion, we show that study type classification in PubMed is opportune, given the available language models. The high accuracy in this study invites extending these models to more elaborate and hierarchical identification schemes.

## Introduction

Systematic reviews and meta-analyses of randomized control trial (RCT) results are the cornerstones of evidence-based medicine. Identifying the relevant RCTs in databases such as PubMed is essential to collecting and collaboratively analyzing new results. PubMed currently offers the largest and most comprehensive collection of abstracts of biomedical studies, currently spanning over 34 million abstracts. Unfortunately, these abstracts are not always accompanied by sufficiently reliable and standardized meta-information (Williamson and Minter 2019). Therefore, to conduct a systematic review, manual screening of all search results is essential to exclude irrelevant publications. This manual screening includes the identification of systematic reviews and meta-analyses that have already been published on the study in question, but also the existing individual RCTs must also be detected. With insufficiently reliable meta-data of study abstracts, this process can go suboptimal in two ways; irrelevant studies may be retrieved, or relevant studies may not be retrieved—the former event results in more reading work, the latter in the unnecessary omission of evidence. Therefore, current search strategies that scan raw abstracts for keywords are consistently set up to achieve high sensitivity (i.e., minimal number of false negatives), but this often results in low specificity (i.e., many false positives) (Beynon et al. 2013; Geersing 2012; Li et al 2019). Therefore, it frequently happens that a systematic review requires thousands of abstracts to be screened to identify all relevant RCTs. In addition to RCTs, PubMed also contains many other study types, including narrative reviews, letters, editorials, preclinical studies, errata, and more fundamental biomedical research. Its low prevalence makes it challenging to identify RCTs without receiving large numbers of false positives. The time-consuming task of identifying relevant literature hampers rapid updating of emerging and often quickly developing evidence. PubMed offers filters for different study types.

Unfortunately, these filters’ sensitivity and specificity are presumably too low to be useful for the identification of previously published systematic reviews, meta-analyses, and RCTs.

We propose a solution based on the rapidly developing field of natural language processing. Instead of creating a filter based on individual words, the overall coherence of the abstract is mapped and linked to a specific study type. General patterns from descriptions of different types of studies – including systematic reviews and RCTs – could then assign abstracts to a specific study type with high sensitivity and specificity. This approach requires a training set of enough abstracts assigned with study-type labels by human reviewers; it also requires proper validation to know whether the classification is sufficiently generalizable to serve the public interest.

In this study, we created a large training set (N=50,000), modeled the overarching patterns, and validated the resulting model on an independent dataset (N=5,000). We compared this new approach to existing PubMed filters. Because the prevalence of systematic reviews and RCTs is relatively low, we tested a new method of creating a training set applying active learning to present all potentially relevant studies to the rater rather than manually classifying all studies individually. Using this approach diminishes raters’ efforts as it allows stopping when these potentially relevant studies are identified rather than labeling all studies in the dataset.

We hypothesized that it would be feasible be possible to generate a classification model, for different types of studies, with high sensitivity and specificity. We also showed a relatively low performance of the internal PubMed filters and an overall efficiency gain in generating a training data set using an active learning approach.

## Methods

### Data collection

In September 2022, we selected two batches of PubMed abstracts. The first dataset was for interrater agreement characterization, model training, and internal validation, and the second dataset was for external validation.

The first batch consists of the first 50,000 abstracts (out of 199,563) in the first week of January 2018 (query: ALL[SB] AND 2018/01/01:2018/01/07[DP] AND HASABSTRACT). This batch was split into an interrater set of 5,000 and a training set of 45,000 abstracts. The second batch consists of the first 5,000 abstracts (out of 255,506) taken four years later (query: ALL[SB] AND 2022/01/01:2022/01/07[DP] AND HASABSTRACT). We deliberately selected our external validation data several years later – after the COVID-19 pandemic – as this would allow us to test the generalizability of our model if the literature gives rise to previously unseen topics and shifts in research trends.

### Data labeling

All abstracts in the interrater set were independently labeled using a pre-specified protocol by two authors with at least fifteen years of PubMed experience (Vinkers and Otte). Abstracts were assigned one of the following study labels: 1) a randomized controlled trial, 2) a human study, 3) a systematic review without meta-analysis, 4) a systematic review with meta-analysis, 5) a study protocol, 6) a rodent study or 7) any other abstract type. Study labels are provided in **Table 1**. Not all abstracts contained valuable texts: in some cases, abstracts referred to a duplicated study. We removed these studies and ended up with a total dataset size of 4,867 abstracts.

**Table 1.** Study labels with pre-specified working definitions used for labeling.

| Study type label | Used working definition |
| --- | --- |
| 1. Randomized controlled trial. | An explicit mention of randomization.<br>Excluding non-human primate and rodent trials. |
| 2. Human study. | Human studies other than randomized controlled trials, observational and intervention (e.g., cohorts, open trials, or case-control studies). |
| 3. Systematic review without meta-analysis. | Excluding non-systematic reviews, non-human studies, protocols, or systematic reviews with a meta-analysis. |
| 4. Systematic review with meta-analysis. | An explicit mention of a meta-analysis. |
| 5. Study protocol. | Study protocol for trials or systematic reviews. |
| 6. Rodent study. | Only primary research studies on rats or mice, excluding rodent cell line research. |
| 7. Misc. | All other studies. |

Labeling was done assisted by active learning. For each study label, the two independent raters selected five relevant and irrelevant abstracts and used this to initiate a Naïve Bayes classifier. The classifier assigned label probabilities to all abstracts and presented the raters with the abstracts that had the highest probabilities for a given class. After each rating, the classifier was updated to re-assign the probabilities. The rating was continued until all studies were identified (operationally decided after 200 presented non-relevant abstracts). Because of the unbalanced distribution of the study types, we chose to use active learning. We used open-source ASReview LAB v1.0 for labeling.^1^

After pooling the active learning results and inventorying all disagreements, we determined the interrater labeling agreement. We relabeled the disagreements independently and resolved the latest remaining label disagreements in a final discussion round. The inter-rater agreement is expressed as Cohen’s kappa coefficient (κ). This qualitative statistic is more robust than simple percent agreement calculation, as it considers the possibility of the agreement occurring by chance. We used the kappa function from the R package ‘psych’ v2.2.9. The training set of 45,000 abstracts was similarly labeled with active learning, except for the prevalent ‘human study’ type. Given the larger dataset and excellent interrater agreement (see Results), the data was split and labeled by one of the two rating authors. We combined this additional dataset with the first 4,867 abstracts and used it for model training.

### Model fine-tuning

We fine-tuned PubMedBERT, a state-of-the-art language model pruned to the PubMed database. PubMedBERT has a transformer architecture to leverage unsupervised pre-training on a large multi-domain corpus of millions of PubMed abstracts by capturing the used scientific and clinical language. The transformer model has learned the textual context by mapping subword relationships in example corpora into a neural network. The model applies an evolving set of mathematical techniques, called attention or self-attention, to detect subtle ways, even distant data elements, in paragraphs up to a few hundred words. This attention mechanism has a substantial advantage compared to previous machine-learning designs. The textual pattern mapping could be done once, as a pre-training, to capture the general structure of a specific text genre effectively. Pre-training is computationally expensive and requires a vast amount of example data. PubMedBERT captures the textual structure of fourteen million PubMed abstracts containing 3.2 billion words (i.e., 21 Gb of raw text data).

We only need to fine-tune this pre-existent language model to our training set for our classification task. To that aim, we added one additional neural network layer to PubMedBERT and trained the entire network on the classification to predict which abstracts belong to which study type. The pre-trained PubMedBERT transformer was downloaded using the HuggingFace Python library (V4.17, python V3.9). The additional network layer had a dropout of 0.2, a reshape of 7, and a SoftMax layer. All abstract texts were tokenized and truncated to a maximum sequence length of 512.

As our set of labels is not larger enough to cover all abstracts – excluding, for example, studies on non-human primates, cell studies, classical reviews, or diagnostic and prognostic studies – we also added an additional study type label: ‘remaining’ to the training set, covering the unlabeled abstracts.

We split the training dataset into train, test, and eval datasets using an 80%, 15%, and 5% split, respectively, and trained the model until the loss did not improve for two epochs (12 epochs total) at a learning rate of 1e^−5^ in batches of 8 abstracts on a V100 NVIDIA graphics processing unit. We tested the model after each epoch (average accuracy on the eval samples) and selected the model instance with the best accuracy for external validation. The model was finally tested on the withheld test dataset (accuracy of 92.45%).

### External validation

The model was trained on 2018 data only. External model validation was done in an independent dataset of 5,000 abstracts sampled in 2022. The two authors (Otte and Vinkers) raters independently labeled a random subset (i.e., 2,500 each) of these abstracts. We characterized the model’s performance with sensitivity and specificity based on the true positive, true negative, false positive, and false negative numbers (human labels considered the ‘gold standard’). Due to the relatively large number of studies not belonging to one of the study type labels, we subsampled this ‘remaining’ category such that we evaluated 1,000 abstracts per rater to the model’s predictions.

### Comparison to PubMed’s internal filter types

PubMed provides filters to select various study types, including ‘Books and Documents’, ‘Clinical Trial’, ‘Meta-Analysis’, ‘Randomized Controlled Trial’, ‘Review’, and ‘Systematic Review’. These labels are assigned to the studies by librarians; however, the exact criteria and overall labeling procedure are not publicly documented. Based on our experience, we notice various false positives and negatives, for example, the ‘Randomized Controlled Trial’ label assigned to an observational cohort study and a systematic review with meta-analysis lacking a ‘Meta-Analysis’ label. Therefore, we quantified the agreement between our human rater dataset (N 4,867) and PubMed’s labels for a subset of labels. We restricted the comparison to ‘Randomized Controlled Trial’, ‘Meta-analysis’ given the lack of definition details on the other categories.

## Results

### Interrater agreement

The active learning approach accelerated the labeling of low-prevalent study types. For example, the prevalence of randomized controlled trials among PubMed abstracts is 4.4%. Systematic reviews with meta-analysis are even scarcer, at 1.7%. The approximate time to read and classify an abstract was 5-10 seconds. The kappa coefficients of the first and second labeling round are presented in **Table 2** and **Table 3**. We found high interrater agreement, particularly if mismatches were reevaluated in an independent second round.

**Table 2.** After the initial active learning round, the interrater agreement is expressed as a kappa coefficient with 95% confidence interval. The total N is based on the finalized version.

| Study type label | Total (N) | Mismatch (N) | $\kappa$ (CI) |
| --- | --- | --- | --- |
| Randomized controlled trial | 218 | 14 | 0.77 (0.65–0.88) |
| Human study | 2267 | 27 | 0.97 (0.96–0.98) |
| Systematic review without meta-analysis | 354 | 32 | 0.78 (0.71–0.85) |
| Systematic review with meta-analysis | 83 | 7 | 0.83 (0.71–0.95) |
| Study protocol | 130 | 4 | 0.90 (0.80–1.00) |
| Rodent study | 403 | 46 | 0.77 (0.71–0.83) |

**Table 3.** After the second active learning round, the interrater agreement is expressed as kappa coefficient with 95% confidence interval. The total N is based on the finalized version.

| Study type label | Total (N) | Mismatch (N) | $\kappa$ (CI) |
| --- | --- | --- | --- |
| Randomized controlled trial | 218 | 2 | 0.97 (0.92–1.00) |
| Human study | 2267 | 9 | 0.99 (0.98–1.00) |
| Systematic review without meta-analysis | 354 | 6 | 0.96 (0.92–0.99) |
| Systematic review with meta-analysis | 83 | 2 | 0.95 (0.88–0.99) |
| Study protocol | 130 | 0 | 1.00 (1.00–1.00) |
| Rodent study | 403 | 7 | 0.97 (0.94–0.99) |

### External validation

We found excellent sensitivity and specificity on the external dataset with abstracts published four years later compared to the training data (**Table 4**). This indicates that the model is generalizable for texts written in different periods (i.e., post-COVID-19 pandemic), as the test dataset dated before the COVID-19 pandemic (2018), whereas the test dataset was four years later (2022).

**Table 4.** The fine-tuned PubMedBERT model’s performance on unseen external data. TP, true positive, TN, true negative, FP, false positive, FN, false negative. CI, 95% confidence interval. We prevented skewed contingency tables and artificially high performances, by pruning the ‘remaining’ category of unlabeled abstracts to a random subset, such that the total sample size per rater equals N 1,000 (originally n=2,500).

| Rater | Study type label | N | TP | TN | FP | FN | Sensitivity | CI<br>(lower) | CI<br>(upper) | Specificity | CI<br>(lower) | CI<br>(upper) |
| --- | --- | --- | --- | --- | --- | --- | --- | --- | --- | --- | --- | --- |
| 1 | Randomized controlled trial | 1000 | 31 | 956 | 5 | 8 | 0.795 | 0.668 | 0.922 | 0.995 | 0.99 | 0.999 |
| 1 | Human study | 1000 | 32 | 796 | 170 | 2 | 0.941 | 0.862 | 1.020 | 0.824 | 0.800 | 0.848 |
| 1 | Systematic review without meta-analysis | 1000 | 25 | 966 | 7 | 2 | 0.926 | 0.827 | 1.025 | 0.993 | 0.987 | 0.998 |
| 1 | Systematic review with meta-analysis | 1000 | 40 | 956 | 4 | 0 | 1.000 | 1.000 | 1.000 | 0.996 | 0.992 | 1.000 |
| 1 | Study protocol | 1000 | 18 | 959 | 23 | 0 | 1.000 | 1.000 | 1.000 | 0.977 | 0.967 | 0.986 |
| 1 | Rodent study | 1000 | 89 | 893 | 9 | 9 | 0.908 | 0.851 | 0.965 | 0.990 | 0.984 | 0.997 |
| 1 | (Remaining, non-labeled) * | 1000 | 542 | 251 | 5 | 202 | 0.728 | 0.697 | 0.760 | 0.980 | 0.964 | 0.997 |
| 2 | Randomized controlled trial | 1000 | 20 | 970 | 8 | 2 | 0.909 | 0.789 | 1.029 | 0.992 | 0.986 | 0.997 |
| 2 | Human study | 1000 | 4 | 799 | 196 | 1 | 0.800 | 0.449 | 1.151 | 0.803 | 0.778 | 0.828 |
| 2 | Systematic review without meta-analysis | 1000 | 11 | 976 | 13 | 0 | 1.000 | 1.000 | 1.000 | 0.987 | 0.980 | 0.994 |
| 2 | Systematic review with meta-analysis | 1000 | 60 | 930 | 9 | 1 | 0.984 | 0.952 | 1.015 | 0.990 | 0.984 | 0.997 |
| 2 | Study protocol | 1000 | 9 | 985 | 6 | 0 | 1.000 | 1.000 | 1.000 | 0.994 | 0.989 | 0.999 |
| 2 | Rodent study | 1000 | 76 | 887 | 35 | 2 | 0.974 | 0.939 | 1.009 | 0.962 | 0.95 | 0.974 |
| 2 | (Remaining, non-labeled) * | 1000 | 551 | 184 | 2 | 263 | 0.677 | 0.645 | 0.709 | 0.989 | 0.974 | 1.004 |

### Comparison to PubMed’s internal filter types

PubMed’s internal ‘Randomized Controlled Trial’ and. ‘Meta-Analysis’ filters only partially captured the ‘gold standard’ manual labels (**Table 5**). The number of false negatives exceeded the number of false positives in both categories. The sensitivity was very low for the ‘Meta-Analysis’ (0.17) and low for the ‘Randomized Controlled Trial’ (0.26), indicating limited usability of PubMed’s internal filters to select, for example, randomized trials for a systematic review.

**Table 5.** PubMed’s internal filter comparison. TP, true positive, TN, true negative, FP, false positive, FN, false negative. CI, 95% confidence interval.

| Study type label | N | TP | TN | FP | FN | Sensitivity | CI<br>(lower) | CI<br>(upper) | Specificity | CI<br>(lower) | CI<br>(upper) |
| --- | --- | --- | --- | --- | --- | --- | --- | --- | --- | --- | --- |
| Systematic review with meta-analysis | 4867 | 7 | 4824 | 3 | 33 | 0.175 | 0.057 | 0.293 | 0.999 | 0.999 | 1.000 |
| Randomized controlled trial | 4867 | 10 | 4820 | 8 | 29 | 0.256 | 0.119 | 0.393 | 0.998 | 0.997 | 0.999 |

### Implementation

We implemented the model in an online cloud platform (https://evidencehunt.com/) after applying the study type prediction to all PubMed abstracts available (34.7 million). Users may query, select and export study types. The database is updated weekly with new abstracts to facilitate scholars and clinicians in need of robust study-type selection and precise search queries of the vast scientific literature.

## Discussion

We show that it is possible to classify study types in PubMed abstracts with high sensitivity and specificity (>95%) based on a transformer model that is fine-tuned on an efficiently constructed training set. Creating a suitable training set requires a lot of time, mainly if the labels include low-prevalence categories. Because RCTs and systematic reviews are relatively infrequent in PubMed, a conventional method of training set construction requires labeling thousands of abstracts. By effectively using active learning, we were able to label only those abstracts that were marked as potentially interesting. This approach confirms a trend in the research field of systematic reviews where active learning has previously been very efficient. The current active learning environment is user-friendly and freely accessible. Because labeling via active learning mostly works with binary classifications (i.e., relevant or irrelevant), we repeated the process for each study type and later combined these labels to create the appropriate dataset for the multiclass classifier. The added value of active learning is interesting for constructing more detailed classification schemas. The added value may be s limited if the prevalence of positive entries is high (as in our case with ‘human study’ type).

One limitation of the current classification algorithm is that it does not cover all existing PubMed abstracts. For example, classical reviews, cell line studies, and prediction studies are missing. Also, our classification only assigns a single label to an abstract. In practice, there are situations where an abstract describes both human and animal results, for example. Open trials are now not separately classified in a separate category, and there is currently no distinction between phase 1, 2, 3, and 4 clinical trials. However, this may be possible in the future with additional training. Hierarchical classification is desirable as meta-analyses may or may not be nested in rodent-oriented or human-oriented systematic reviews, but also necessary to establish a classification scheme separating children and adults. RCTs are possible in children and adults, so assigning multiple labels to a single abstract is a necessary next step.

Machine learning models require exceedingly larger training data sets, we show that using active learning can accelerate the labeling process. With the exponential increase of publications each year, finding relevant publications is becoming more difficult. Here we show that transformer models can aid in selecting studies, potentially preventing wasting precious time and energy to find relevant publications.

## Data Availability

All data produced in the present study are available upon reasonable request to the authors

https://evidencehunt.com/

## Footnotes

1 https://asreview.nl/

